# Achieving Expert-Level Clinical Infection Detection with LLMs from Clinical Documents: Validation in Complex Patient Cases with Cirrhosis

**DOI:** 10.64898/2026.01.13.26344046

**Authors:** Yufei Yu, James Ford, Eileen Kim, Avi Patel, Gabriel Wardi, Rohit Loomba, Atul Malhotra, Shamim Nemati, Joseph C. Ahn

## Abstract

**Background:** Systemic infections are a leading cause of hospitalization and death among patients with cirrhosis. Timely and accurate infection identification is essential for both clinical care and the development of predictive models. However, existing methods such as ICD-10 coding are unreliable, and manual chart review is resource-intensive and difficult to scale. This study aimed to develop and validate an automated large language model (LLM)-based approach for infection classification and subtyping in patients with cirrhosis presenting to the emergency department (ED).

**Method:** We developed INFEHR (INfection identification and subtyping using Free-text EHR analysis), an LLM-powered pipeline utilizing Claude 3.5 Sonnet to analyze clinical notes from the first 72 hours of admission. Model outputs were compared against a physician-adjudicated gold standard in a cohort of 1,000 encounters from patients with cirrhosis who presented to the ED. Performance was benchmarked against ICD-10 code–based labeling and CDC Adult Sepsis Event criteria.

**Results:** INFEHR achieved 94.7% overall accuracy, with 99.5% sensitivity and 92.8% positive predictive value for identifying infection presence, outperforming ICD-10–based classification across all metrics (*p* < 0.0001). The model also demonstrated strong performance in classifying pathogen type and infection site. This pipeline processed notes within seconds, offering improvements in efficiency and scalability over manual review.

**Conclusion:** INFEHR offers a scalable, reproducible, and accurate method for infection phenotyping in cirrhosis. By overcoming limitations of traditional coding and manual review, it supports high-throughput infection surveillance, improves cohort construction for clinical research, and enables future integration into real-time decision-support tools in hepatology.

## 1. INTRODUCTION

Cirrhosis, the end-stage of chronic liver disease, affects over 5 million people globally and causes nearly 1.5 million annual deaths^1^. It leads to systemic complications, including immune dysfunction that increases infection risk^2, 3^; 25–47% of hospitalized cirrhotic patients develop bacterial infections^4^, a rate far exceeding that of the general hospital population. These infections often precipitate serious complications with high mortality, like variceal hemorrhage, hepatic encephalopathy, hepatorenal syndrome, and acute-on-chronic liver failure^4, 5^.

Diagnosing infections in cirrhosis is challenging. Systemic inflammation can mimic sepsis; up to 30% of decompensated cirrhosis cases meet systemic inflammatory response syndrome (SIRS) criteria despite no infection^5^. Conversely, true infections in cirrhosis may have atypical presentations due to cirrhosis-associated immune paralysis, leading standard sepsis tools to misclassify patients^5^. This can delay treatment^6^ or prompt unnecessary antimicrobial use, increasing resistance risk^7, 8^. These diagnostic complexities demand improved tools for timely and accurate infection recognition, as treatment delays significantly worsen outcomes^9^. Yet, no validated criteria or decision-support systems currently exist to guide infection risk stratification or early antimicrobial initiation in this high-risk population.

Identifying infection type and anatomical source(e.g., pneumonia, urinary tract infection) is essential for clinical care, predictive modeling, and epidemiologic research. Ground truth labels are foundational to these efforts, but expert chart review is labor-intensive and unscalable^10^. Surrogate methods like International Classification of Diseases (ICD) codes suffer from misclassification, omissions, and variability due to billing-driven practices^11–13^. Rule-based tools like the Linder-Mellhammar Criteria for Infection^14^ also underperform in cirrhosis, where overlapping inflammatory responses and atypical presentations lead to high false-positive rates.

Recent advances in natural language processing, particularly large language models (LLMs), offer new potential for extracting complex information from unstructured electronic health record (EHR) text^15–21^. LLMs have shown promise across infection-related tasks, including sepsis detection^22^, surgical site infection classification^23^, bloodstream infection identification^24, 25^, and antimicrobial resistance extraction^26^. These approaches often match or outperform that of traditional methods^21, 23–27^, yet many prior studies are often small and lack statistical power^23–26^, limiting generalizability. Nonetheless, these findings suggest LLMs can perform nuanced clinical reasoning needed for infection identification in complex cases.

In this study, we proposed and evaluated an automated LLM-based chart review pipeline for infection classification in patients with cirrhosis during the first 72 hours following admission. We compared its performance to expert manual chart review, aiming to generate accurate infection labels at scale, offering greater efficiency than manual review and improved precision over administrative codes. This work represents the largest evaluation of automated infection classification in cirrhosis, both in cohort size and granularity of infection subtyping. The focus on cirrhosis, a population with complex and atypical infection presentations, underscores the clinical value of this approach where traditional methods often underperform.

## 2. METHODS

### 2.1. Pipeline Overview

Figure 1 presents an overview of INFEHR (**IN**fection identification and subtyping using **F**ree-text **EHR** analysis), an LLM-based chart review pipeline for infection classification during the initial days of admission in cirrhotic patients. For each patient encounter, clinical notes from the first 72 hours following admission were extracted. This time period was intended to capture early diagnostic signals relevant for time-sensitive tasks such as antibiotic de-escalation. These notes were submitted to an LLM, which generated a structured output for infection presence, pathogen type, and infection location.

**Figure 1.**
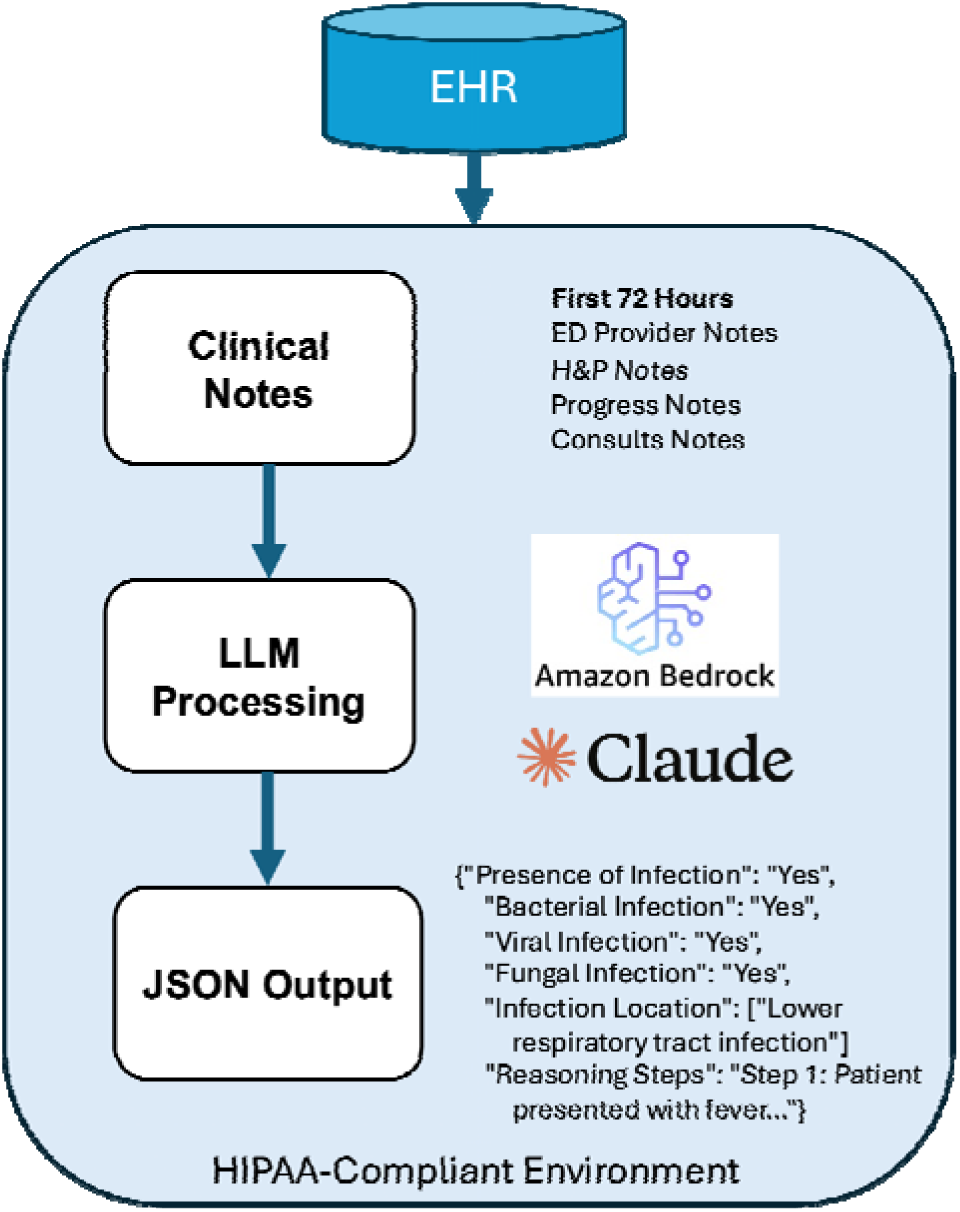
Overview of the INFEHR (**I**nfection ide**N**tification and subtyping using **F**ree-text **EHR** analysis) pipeline. Clinical notes (ED provider notes, H&P notes, Progress notes, and Consult notes) from the first 72 hours of a patient’s encounter are processed by an LLM within a HIPAA-compliant environment. The pipeline outputs structured data in JSON format, identifying key clinical concepts such as the presence and type of infection, location, and the reasoning derived from the text.

### 2.2. Study Setting and Participants

This study included adult patients with cirrhosis, identified using specific ICD-10 codes (K70.3, K70.31, K71.7, K74.3, K74.4, K74.5, K74.6, K74.69), who presented to the emergency department (ED) at UC San Diego Health between 2016 and 2023. Eligible encounters included patients who were either admitted or placed in observation status following their ED visit. Encounters with a total length of stay less than 48 hours were excluded to ensure sufficient documentation, as infections in cirrhotic patients often require prolonged care and bacterial culture results may take several days to return. This investigation was conducted according to the UCSD Institutional Review Boards (IRB) approved protocol #805726 with a waiver of informed consent.

### 2.3. Data Processing

Data were extracted from the Epic Clarity database and securely stored in a HIPAA-compliant environment. For each encounter, the first 72 hours of ED provider, history and physical (H&P), progress, and consultation notes were extracted and concatenated chronologically to preserve the temporal sequence of clinical documentation for LLM input.

### 2.4. Large Language Model Prompting and Output

To extract infection-related information from clinical notes, we utilized Anthropic’s Claude 3.5 Sonnet^28^ to process the complete 72-hour note set for each encounter. The model was guided by a prompt that simulated expert clinical reasoning to evaluate infection presence, pathogen type, and anatomical site based on clinical documentation. At each step, the model reported a confidence level – strong, intermediate, or low – reflecting its self-assessed certainty based on the available evidence.

The model first assessed for acute infection using clinical indicators such as vital signs, laboratory results, imaging findings, microbiology, and provider documentation, excluding chronic infections like HIV or hepatitis C. If infection was identified, it then classified the pathogen type (bacterial, viral, or fungal) and, when possible, the anatomical source using predefined categories, including lower respiratory tract, urinary tract, intra-abdominal, gastrointestinal, skin and soft tissue, bone and joint, central nervous system (CNS), primary bloodstream, catheter-related, neutropenic fever, ear, nose, throat (ENT), and reproductive tract infections^14^. To avoid overclassification, the model was explicitly directed not to rely solely on empiric or prophylactic antibiotic use.

To support consistency and interpretability, the model generated structured outputs containing binary classifications for infection presence and pathogen type, identified infection sites, and brief justifications. It also produced a reasoning array mimicking clinical logic by 1) summarizing key points; 2) correlating findings with diagnostic data; and 3) explaining the final classification. The complete prompt is provided in **Supplementary Material 2**, with detailed prompt design described in **Supplementary Methods Section 1**.

### 2.5. Cohort Selection

We selected two cohorts to develop and evaluate INFEHR (Figure 2). There was no overlap between the prompt development and evaluation cohorts.

**Figure 2.**
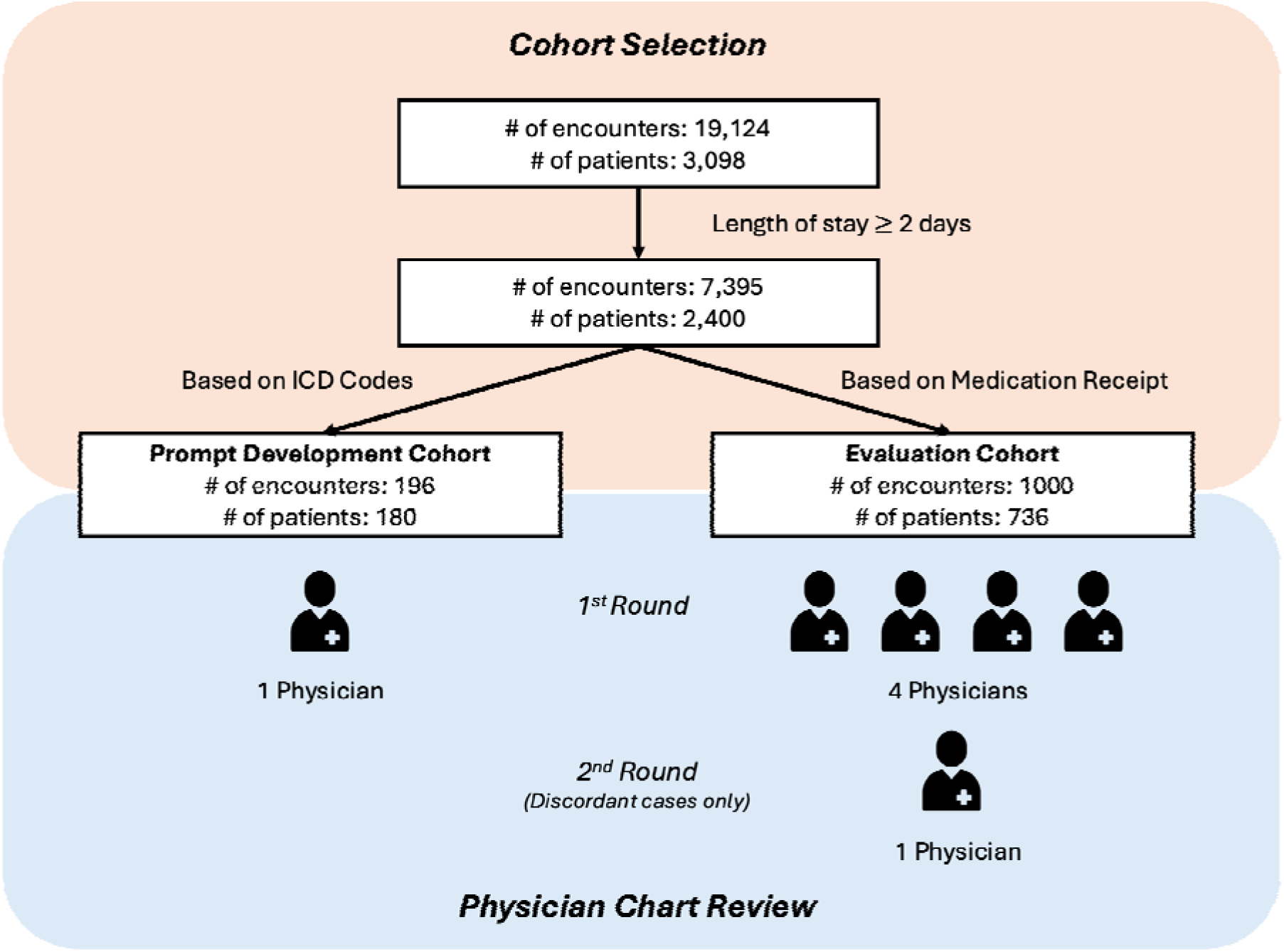
Cohort selection and chart review process. Patient cohorts were identified from initial EHR encounters. Inclusion criteria, including length of stay, medication administration records, and ICD codes, were applied to define the prompt development and evaluation cohorts. Manual chart review by physicians (1 for prompt development, 4 for evaluation) established ground truth labels.

The prompt development cohort was used to refine INFEHR’s prompt and logic. This cohort was selected using ICD-10 codes associated with the hospitalization, followed by manual review of discharge summaries to ensure broad representation, including less common infection types and locations. Full performance metrics analysis for the prompt development cohort is available in **Supplementary Results Section 1**.

Once INFEHR was refined, its performance was tested using the evaluation cohort and compared against physician chart review (gold standard). To ensure adequate representation of different scenarios and mitigate potential biases associated with ICD code selection alone, this cohort was identified based on documented administration of anti-infective medications during hospitalization. Encounters were sampled to achieve the following distribution: 500 with antibiotics, 100 with antivirals, 50 with antifungals, and 350 with no anti-infective medications administered.

### 2.6. Performance Evaluation

Gold-standard labels for the prompt development cohort were established through chart review by an expert physician. Labels were generated in three domains and used to assess model performance against INFEHR outputs:

1. Infection presence (infection vs. no infection)
2. Pathogen type (bacterial, viral, fungal)
3. Infection location (12 predefined categories)

For the evaluation cohort, chart review was independently conducted by four physicians using the same three classification domains and the same 72-hours input window as the INFEHR pipeline. The reviewers included one specialist in gastroenterology and hepatology (J.A.), two emergency medicine physicians (J.F. and A.P.), and one internal medicine physician (E.K.). Each reviewer provided infection classifications along with a binary confidence rating (yes/no) for each classification decision, indicating their certainty based on the available clinical documentation. To assess inter-rater agreement, Fleiss’ kappa was calculated between three reviewers using a subset of 100 encounters. Cases with disagreement between the initial physician review and INFEHR’s output on infection presence were adjudicated by a single physician (J.A.) who was blinded to the model output. As a baseline comparison, infection classification performance using ICD-10 codes recorded during the encounter was evaluated. The list of ICD-10 codes was curated by two physicians (**Supplementary Table 1**). Additionally, INFEHR’s performance for infection presence was compared against the CDC’s Adult Sepsis Event definition of presumed serious infection, defined as a blood culture order and administration of at least four days of intravenous antibiotics^29^.

Pipeline performance was evaluated using standard classification metrics: accuracy, sensitivity, specificity, positive predictive value (PPV), and negative predictive value (NPV). Statistical comparisons between INFEHR and ICD-10 codes for accuracy, sensitivity, and specificity were performed using McNemar’s test for paired nominal data. Comparisons of PPV and NPV utilized the relative value method described by Moskowitz and Pepe^30, 31^. INFEHR confidence levels were converted to a numeric scale (1=Low, 2=Intermediate, 3=Strong) for quantitative analysis.

### 2.7. Computational Implementation

INFEHR was deployed in a HIPAA-compliant Amazon Web Services (AWS) instance^32^, configured with authentication and permissions to securely interact with Claude 3.5 Sonnet v2 through Amazon Bedrock^33^. To support efficient scaling, the system could be configured to enable parallel processing (e.g., via AWS Lambda)^34^, allowing multiple encounters to be reviewed simultaneously.

## 3. RESULTS

### 3.1. Study Cohorts

We identified 7,395 ED encounters from 2,400 unique adult patients with cirrhosis between 2016 and 2023 at UCSD Health. Patients were included if they had cirrhosis-related ICD-10 codes and a length of stay ≥ 48 hours. The full cohort derivation is shown in Figure 2.

The evaluation cohort comprised 1,000 encounters and underwent manual chart review by four physicians. Inter-rater agreement for infection presence was substantial (Fleiss’ kappa = 0.704, 95% CI: 0.590–0.812). For pathogen and site classification, the mean Jaccard indices were 0.728 (95% CI: 0.662–0.793) and 0.671 (95% CI: 0.594–0.740), respectively. 59.7% of encounters (n=597) involved a confirmed infection by chart review. Among these, 557 encounters received antibiotics, 198 received antivirals, and 72 received antifungals. Physician reviewers expressed confidence in their infection presence classification for 80.7% of cases. Additional cohort characteristics are summarized in **Supplementary Table 2**.

### 3.2. Infection Presence Identification Performance

In the initial evaluation of 1,000 encounters, INFEHR achieved 86.5% accuracy, with a sensitivity of 96.5%, specificity of 71.7%, PPV of 83.5%, and NPV of 93.2%. There were 135 discordant cases between the model and physician reviewers. These were independently adjudicated by a physician who was blinded to the model’s outputs. INFEHR’s classification was confirmed in 60.0% (n=81) of these cases. Using the adjudicated results as the reference standard, INFEHR’s accuracy improved to 94.7%, with a sensitivity of 99.5%, specificity of 86.0%, PPV of 92.8%, and NPV of 99.0% (**Table 1**).

**Table 1.** Performance metrics for infection presence identification in the evaluation cohort: INFEHR pipeline vs. ICD-10 codes. Statistical comparisons were conducted between INFEHR and each baseline method.

| Methods | Accuracy | Sensitivity | Specificity | PPV | NPV |
| --- | --- | --- | --- | --- | --- |
| INFEHR | 0.947 | 0.995 | 0.860 | 0.928 | 0.990 |
| ICD-10 Codes | 0.752**** | 0.843**** | 0.588**** | 0.787**** | 0.675**** |
| CDC<br>Presumed<br>Serious<br>Infection | 0.784**** | 0.840**** | 0.683**** | 0.827**** | 0.703**** |

\*\*\*\* represents p-value $\leq 0.0001$ .

INFEHR’s confidence scores reflected the strength of supporting clinical evidence. It assigned a strong confidence rating in 90.7% (n=907) of cases, and intermediate confidence in the remaining 9.3% (n=93). The model was more confident when classifying encounters as infection-negative than infection-positive (p < 0.0001).

INFEHR was reliable when expressing high confidence. In discordant cases where the model expressed strong confidence but the reviewer did not, the model’s classification aligned with the adjudicated label nearly 80% of the time. Even when both the model and reviewer had low or intermediate confidence, the model’s classification was favored in roughly half of cases (**Supplementary Table 3**).

Compared to INFEHR (**Table 1**), ICD-10 code–based classification showed lower performance across all metrics (p < 0.0001), and similarly underperformed when using the CDC’s definition of presumed serious infection (p < 0.0001).

### 3.3. Pathogen Type Performance

In the evaluation cohort, bacterial infections were present in 58.9% (n=589), viral in 5.7% (n=57), and fungal in 3.8% (n=38), based on the final adjudicated standard. INFEHR demonstrated high sensitivity across all pathogen types, and outperformed ICD-10-based classification particularly in sensitivity for viral and fungal types (**Table 2**).

**Table 2.**
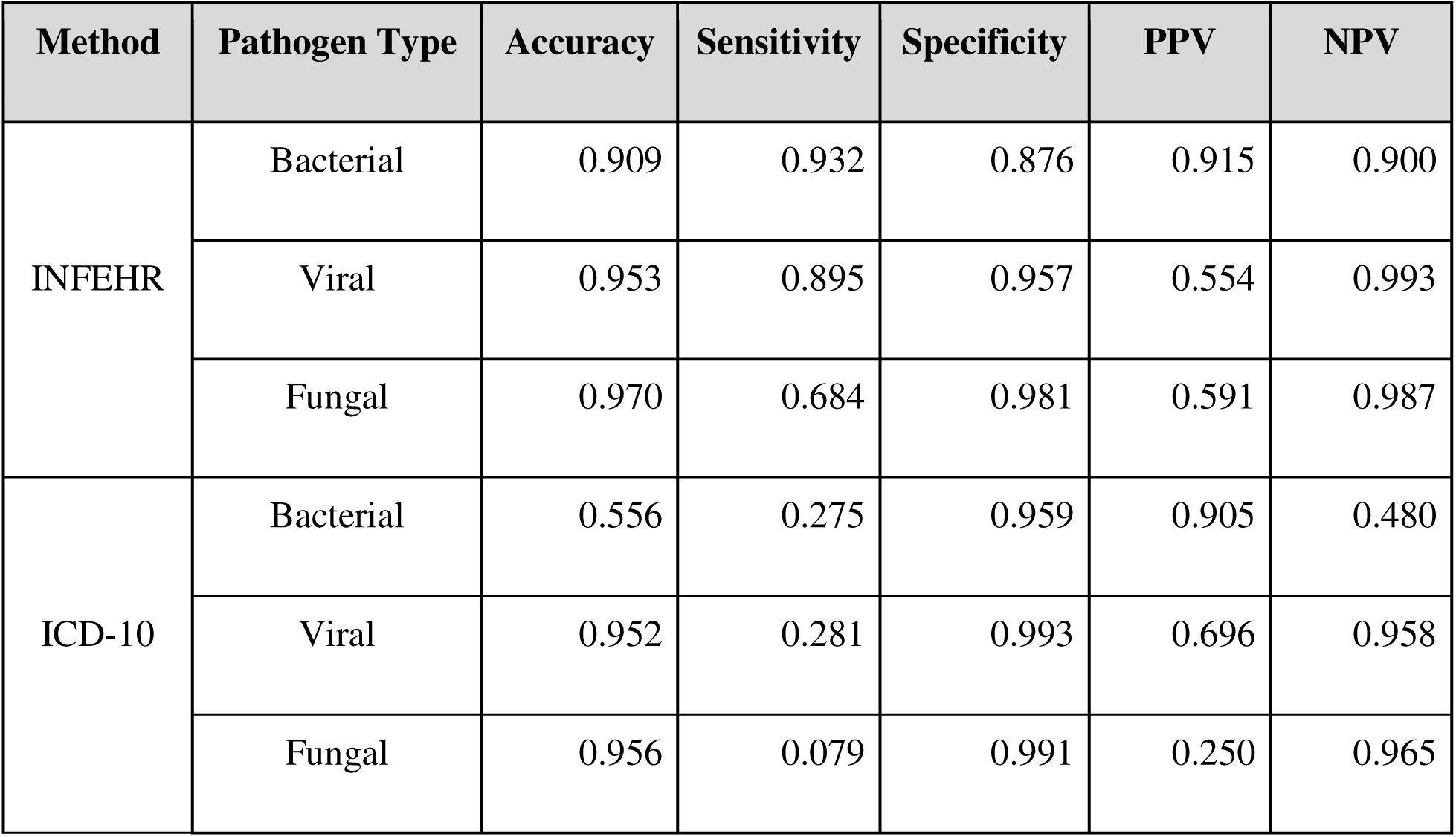
Performance metrics for pathogen type classification in the evaluation cohort: INFEHR pipeline vs. ICD-10 codes.

Model-assigned confidence scores on a 3-point scale averaged 2.76 for bacterial infections and 2.97 for both viral and fungal infections. As with infection presence, INFEHR tended to assign lower confidence to positive classifications (e.g., 2.66 for bacterial-positive cases) and higher confidence to negatives (e.g., 2.91 for bacterial-negative cases), reflecting greater certainty when ruling out infection.

### 3.4. Infection Location Performance

Among infections confirmed in the adjudicated gold standard, the most common sites were lower respiratory tract (n=154), intra-abdominal (n=145), skin and soft tissue (n=122), primary bloodstream (n=110), and urinary tract (n=108). Less frequent sites included gastrointestinal (n=71), bone and joint (n=21), ENT (n=10), CNS (n=8), catheter-related (n=5), neutropenic fever (n=4), and reproductive tract (n=1).

INFEHR demonstrated strong performance in identifying common infection sites (**Table 3**). Sensitivity and PPV were highest for urinary tract, skin and soft tissue, and lower respiratory tract infections. For rarer infections such as CNS, catheter-related, and reproductive tract, performance was limited. Compared to ICD-10–based classification (**Supplementary Table 6**), INFEHR consistently showed better sensitivity across all infection sites, with substantial gains for common infections like lower respiratory tract or intra-abdominal infection. While ICD-10 codes yielded very high specificity (>99%), they often missed true positives.

**Table 3.** INFEHR performance for infection location classification in the evaluation cohort.

| Infection Location | TP | TN | FP | FN | Accuracy | Sensitivity | Specificity | PPV | NPV | Confidence |
| --- | --- | --- | --- | --- | --- | --- | --- | --- | --- | --- |
| Lower Respiratory Tract | 128 | 805 | 41 | 26 | 0.933 | 0.831 | 0.952 | 0.757 | 0.969 | 2.27 |
| Urinary Tract | 94 | 865 | 27 | 14 | 0.959 | 0.870 | 0.970 | 0.777 | 0.984 | 2.74 |
| Intra-abdominal | 108 | 823 | 32 | 37 | 0.931 | 0.744 | 0.963 | 0.771 | 0.957 | 2.66 |
| Gastrointestinal | 43 | 912 | 17 | 28 | 0.955 | 0.606 | 0.982 | 0.717 | 0.970 | 2.70 |
| Skin and Soft Tissue | 106 | 855 | 23 | 16 | 0.961 | 0.869 | 0.974 | 0.822 | 0.982 | 2.74 |
| Bone and Joint | 18 | 975 | 4 | 3 | 0.993 | 0.857 | 0.996 | 0.818 | 0.997 | 2.82 |
| Central Nervous System | 4 | 989 | 3 | 4 | 0.993 | 0.500 | 0.997 | 0.571 | 0.996 | 2.14 |
| Primary Bloodstream | 77 | 853 | 37 | 33 | 0.930 | 0.700 | 0.958 | 0.675 | 0.963 | 2.74 |
| Catheter-related | 5 | 989 | 6 | 0 | 0.994 | 1.000 | 0.994 | 0.455 | 1.000 | 2.73 |
| Neutropenic Fever | 3 | 995 | 1 | 1 | 0.998 | 0.750 | 0.999 | 0.750 | 0.999 | 3.00 |
| Ear, Nose, Throat | 6 | 981 | 9 | 4 | 0.987 | 0.600 | 0.991 | 0.400 | 0.996 | 2.60 |
| Reproductive Tract | 0 | 998 | 1 | 1 | 0.998 | 0.000 | 0.999 | 0.000 | 0.999 | 3.00 |

### 3.5. Scalability and Efficiency

INFEHR processed each encounter in an average of 16.5 seconds. Leveraging parallel processing through AWS Lambda, which supports up to 1,000 simultaneous executions by default, the system can theoretically process 1,000 encounters in approximately the same amount of time, 16.5 seconds. In contrast, manual chart review by physicians required at least 2 minutes per encounter, translating to over 30 person-hours to review 1,000 encounters (Figure 3).

**Figure 3.**
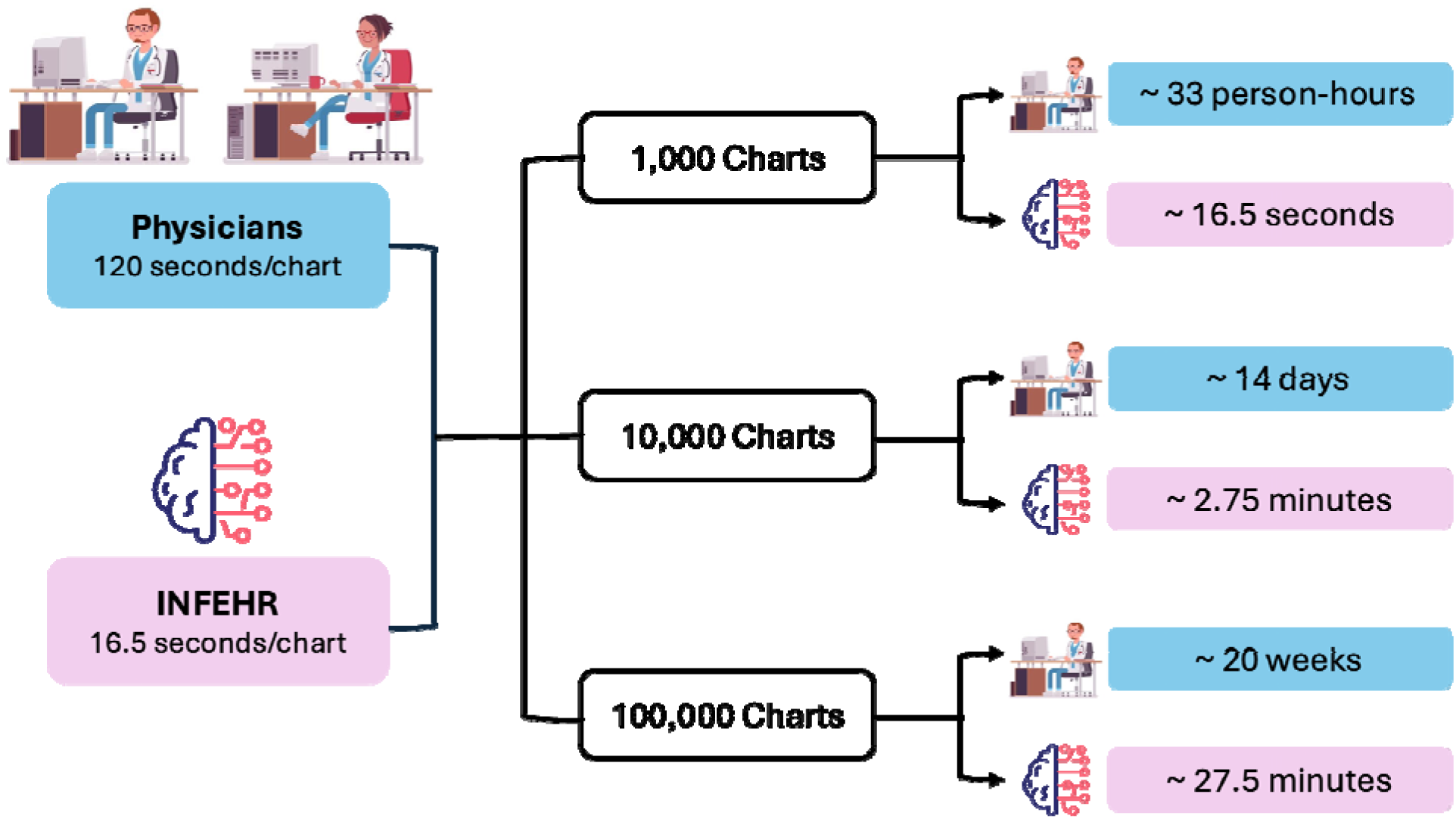
Time efficiency comparison of manual review vs. INFEHR pipeline at scale. Estimated manual physician review required an average of 120 seconds per clinical chart, whereas the INFEHR pipeline averaged 16.5 seconds per chart. The total time for INFEHR accounts for concurrent processing capabilities (e.g., 1,000 concurrent executions via Amazon Lambda).

## 4. DISCUSSION

This study demonstrates that INFEHR, an LLM-powered pipeline, can accurately classify infection presence, pathogen type, and infection site from free-text clinical documentation in the early phase of admission, specifically in patients with cirrhosis, one of the most diagnostically challenging populations. INFEHR achieved high concordance with expert physician chart review and outperformed both ICD-10-based classification and CDC-defined criteria for presumed serious infection. In discordant cases, secondary adjudication more frequently favored the model over initial human reviews, highlighting its potential to augment diagnostic accuracy. Beyond accuracy, INFEHR improves both scalability and efficiency. It processes full clinical notes within seconds per encounter, reducing the labor-intensive demands of manual chart review. Additionally, by applying consistent decision criteria across all encounters, INFEHR reduces inter-rater variability and enhances reproducibility.

Compared to traditional approaches, INFEHR addresses known limitations of traditional infection phenotyping in cirrhosis. ICD-10 codes, frequently used in retrospective research, are prone to undercapture infections due to delayed assignment, billing-oriented abstraction, and lack of clinical specificity^13, 35–39^. They are ill-suited for capturing time-sensitive information, such as identifying infections at or near the time of admission. Similarly, the CDC’s Adult Sepsis Event definition of a “presumed serious infection” also does not reliably capture the true onset of infection^40^ and may misclassify infection status in cirrhotic patients. Empiric antibiotics are routinely initiated during episodes of ascites, hepatic encephalopathy, or gastrointestinal bleeding, leading to overestimation of true infection prevalence^41, 42^. In contrast, INFEHR provides a more tailored and accurate approach, capable of distinguishing true infections from precautionary or prophylactic treatments by leveraging nuanced clinical documentation.

Discrepancies between INFEHR and physician reviewers often arose in cases with ambiguous documentation, empiric therapies, or borderline presentations, areas where diagnostic uncertainty is common even among experts. In such instances, establishing a definitive ground truth is inherently challenging. Notably, adjudication sided with INFEHR in 60% of discordant cases, indicating that the model successfully identified clinically meaningful infections that may have been overlooked or inconsistently documented in the initial chart review. Human factors such as variation in clinical training, specialty background, and cognitive fatigue from assessing hundreds of encounters may contribute to labeling inconsistencies, reinforcing the limitations of manual review as a gold standard. Consequently, our results may be biased toward the null^43^, potentially underestimating INFEHR’s true diagnostic performance.

While INFEHR demonstrates strong performance in accurately classifying straightforward cases, it is not intended to replace expert clinical judgment. Human review remains indispensable for complex presentations, particularly in patients with multiple comorbidities, atypical presentations, or conflicting documentation. INFEHR’s internal confidence scores were highly correlated with diagnostic accuracy, suggesting their potential utility for guiding workflow decisions. A hybrid “model-in-the-loop” approach could optimize both efficiency and diagnostic reliability: high-confidence classifications from INFEHR are accepted automatically, while lower-confidence cases are flagged for expert review. This approach may increase the efficiency of infection phenotyping while maintaining diagnostic rigor, especially for large-scale research or surveillance efforts.

INFEHR provides a scalable, validated tool for infection phenotyping in patients with cirrhosis, which has broad implications for epidemiological surveillance and clinical research. By accurately identify pathogen class (i.e. bacterial) and infection site (i.e., lower respiratory tract), INFEHR enables more granular risk stratification and supports robust studies on infection-related outcomes. These structured outputs facilitate the creation of silver-standard datasets that can be leveraged for training and validating predictive models^44^, helping to overcome the major bottleneck of manual chart annotation. By enabling high-quality, automated labeling with demonstrated concordance to expert chart review, INFEHR enhances the quality and feasibility of secondary EHR data analyses in cirrhosis research.

This study has limitations. It was conducted at a single large academic medical center, which may limit generalizability to other healthcare systems. INFEHR’s performance varied across rare subtypes and locations (e.g., fungal infections, reproductive tract infections), likely due to small sample sizes in these subsets. Like human reviewers, the model’s accuracy is contingent on the clarity and completeness of clinical documentation, an inherent challenge in real-world EHR data. Our analysis was also confined to the first 72 hours of admission and a select set of note types. Expanding the temporal window (i.e., >72 hours after admission) and incorporating additional note types could alter model performance and should be explored in future work. While hallucinations were rare, LLMs remain vulnerable to generating factually inconsistent outputs, underscoring the need for safeguards and human oversight in deployment. Lastly, this study evaluated only Anthropic’s Claude 3.5 Sonnet; broader benchmarking across alternative LLMs is necessary to assess generalizability and optimize performance across diverse clinical environments.

In conclusion, our LLM-based approach INFEHR achieved high accuracy in determining infection presence and classifying pathogen types, superior to traditional ICD-10 code methods, offering a highly efficient, scalable alternative to manual chart review. This work highlights how LLMs can enhance clinical research efficiency by reliably processing complex, unstructured text. Beyond retrospective analysis, the ability to extract standardized infection labels at scale opens new opportunities for developing reliable real-time clinical decision-support tools.

## ACKNOWLEDGEMENTS

Y.Y. is funded by the U.S. National Library of Medicine (T15LM011271). S.N. is funded by the U.S. National Institute of General Medical Sciences (R35GM143121). J.C.A. is funded by the NIH AIM-AHEAD Program Agreement NO. 1OT2OD032581 and AASLD CTORA25-167853. The authors would like to thank Dr. Eliah Aronoff-Spencer for his feedback and support during the preliminary phase of the study.

## AUTHOR CONTRIBUTION

Y.Y., S.N., and J.C.A. were involved in the original conception and design of the work. Y.Y. developed the pipeline, conducted the experiments, and analyzed the data. J.F., E.K., A.P., and J.C.A. provided clinical expertise and reviewed patient data. Y.Y. and J.C.A. contributed to interpretation of results. All authors contributed to manuscript preparation, critical revisions, and have read and approved the manuscript.

## DATA AVAILABILITY

Access to the de-identified UCSD cohort can be made available by contacting the corresponding author and via approval from the UCSD Institutional Review Boards (IRB) and Health Data Oversight Committee (HDOC).

## CODE AVAILABILITY

The code for computing performance metrics is available at https://github.com/NematiLab/INFEHR.

## REFERENCES

1. Liu YB, Chen MK. Epidemiology of liver cirrhosis and associated complications: Current knowledge and future directions. World J Gastroenterol 2022;28:5910–5930.

2. Bonnel AR, Bunchorntavakul C, Reddy KR. Immune dysfunction and infections in patients with cirrhosis. Clin Gastroenterol Hepatol 2011;9:727–38.

3. Hasa E, Hartmann P, Schnabl B. Liver cirrhosis and immune dysfunction. Int Immunol 2022;34:455–466.

4. Bruns T, Zimmermann HW, Stallmach A. Risk factors and outcome of bacterial infections in cirrhosis. World J Gastroenterol 2014;20:2542–54.

5. Miranda-Zazueta G, Leon-Garduno LAP, Aguirre-Valadez J, et al. Bacterial infections in cirrhosis: Current treatment. Ann Hepatol 2020;19:238–244.

6. Piano S, Tonon M, Angeli P. Changes in the epidemiology and management of bacterial infections in cirrhosis. Clin Mol Hepatol 2021;27:437–445.

7. Llor C, Bjerrum L. Antimicrobial resistance: risk associated with antibiotic overuse and initiatives to reduce the problem. Ther Adv Drug Saf 2014;5:229–41.

8. Shallcross LJ, Davies DS. Antibiotic overuse: a key driver of antimicrobial resistance. Br J Gen Pract 2014;64:604–5.

9. Arabi YM, Dara SI, Memish Z, et al. Antimicrobial therapeutic determinants of outcomes from septic shock among patients with cirrhosis. Hepatology 2012;56:2305–15.

10. Henry KE, Hager DN, Osborn TM, et al. Comparison of Automated Sepsis Identification Methods and Electronic Health Record-based Sepsis Phenotyping: Improving Case Identification Accuracy by Accounting for Confounding Comorbid Conditions. Crit Care Explor 2019;1:e0053.

11. Glenn DA, Zee J, Hegde A, et al. Validation of Diagnosis Codes to Identify Infection-Related Acute Care Events in Patients With Glomerular Disease. Kidney Int Rep 2021;6:3079–3082.

12. Jolley RJ, Sawka KJ, Yergens DW, et al. Validity of administrative data in recording sepsis: a systematic review. Crit Care 2015;19:139.

13. Karlic KJ, Clouse TL, Hogan CK, et al. Comparison of Administrative versus Electronic Health Record-based Methods for Identifying Sepsis Hospitalizations. Ann Am Thorac Soc 2023;20:1309–1315.

14. Mellhammar L, Elen S, Ehrhard S, et al. New, Useful Criteria for Assessing the Evidence of Infection in Sepsis Research. Crit Care Explor 2022;4:e0697.

15. Agbareia R, Omar M, Soffer S, et al. Visual-textual integration in LLMs for medical diagnosis: A preliminary quantitative analysis. Comput Struct Biotechnol J 2025;27:184–189.

16. Boussina A, Krishnamoorthy R, Quintero K, et al. Large Language Models for More Efficient Reporting of Hospital Quality Measures. NEJM AI 2024;1.

17. Feng R, Brennan KA, Azizi Z, et al. Engineering of Generative Artificial Intelligence and Natural Language Processing Models to Accurately Identify Arrhythmia Recurrence. Circ Arrhythm Electrophysiol 2025;18:e013023.

18. Ntinopoulos V, Rodriguez Cetina Biefer H, Tudorache I, et al. Large language models for data extraction from unstructured and semi-structured electronic health records: a multiple model performance evaluation. BMJ Health Care Inform 2025;32.

19. Singhal K, Azizi S, Tu T, et al. Large language models encode clinical knowledge. Nature 2023;620:172–180.

20. Van Veen D, Van Uden C, Blankemeier L, et al. Clinical Text Summarization: Adapting Large Language Models Can Outperform Human Experts. Res Sq 2023.

21. Yuan K, Yoon CH, Gu Q, et al. Transformers and large language models are efficient feature extractors for electronic health record studies. Commun Med (Lond) 2025;5:83.

22. Shashikumar SP, Mohammadi S, Krishnamoorthy R, et al. Development and Prospective Implementation of a Large Language Model based System for Early Sepsis Prediction. NPJ Digit Med 2025.

23. Badia JM, Casanova-Portoles D, Membrilla E, et al. Evaluation of ChatGPT-4 for the detection of surgical site infections from electronic health records after colorectal surgery: A pilot diagnostic accuracy study. J Infect Public Health 2025;18:102627.

24. Perret J, Schmid A. Application of OpenAI GPT-4 for the retrospective detection of catheter-associated urinary tract infections in a fictitious and curated patient data set. Infect Control Hosp Epidemiol 2024;45:96–99.

25. Rodriguez-Nava G, Egoryan G, Goodman KE, et al. Performance of a large language model for identifying central line-associated bloodstream infections (CLABSI) using real clinical notes. Infect Control Hosp Epidemiol 2024;46:1–4.

26. Lorenzoni G, Garbin A, Brigiari G, et al. Large Language Models in Action: Supporting Clinical Evaluation in an Infectious Disease Unit. Healthcare (Basel) 2025;13.

27. Iscoe M, Socrates V, Gilson A, et al. Identifying signs and symptoms of urinary tract infection from emergency department clinical notes using large language models. Acad Emerg Med 2024;31:599–610.

28. Claude 3.5 Sonnet. Volume 2025: Anthropic.

29. Rhee C, Dantes RB, Epstein L, et al. Using objective clinical data to track progress on preventing and treating sepsis: CDC’s new ‘Adult Sepsis Event’ surveillance strategy. BMJ Qual Saf 2019;28:305–309.

30. Moskowitz CS, Pepe MS. Comparing the predictive values of diagnostic tests: sample size and analysis for paired study designs. Clin Trials 2006;3:272–9.

31. Park C, Park SY, Kim HJ, et al. Statistical Methods for Comparing Predictive Values in Medical Diagnosis. Korean J Radiol 2024;25:656–661.

32. Compute Service - Amazon EC2. Volume 2025: Amazon Web Service.

33. Amazon Bedrock. Volume 2025: Amazon Web Service.

34. Understanding Lambda function scaling. AWS Lambda. Volume 2025: Amazon Web Service.

35. Higgins TL, Deshpande A, Zilberberg MD, et al. Assessment of the Accuracy of Using ICD-9 Diagnosis Codes to Identify Pneumonia Etiology in Patients Hospitalized With Pneumonia. JAMA Netw Open 2020;3:e207750.

36. Khokhar B, Jette N, Metcalfe A, et al. Systematic review of validated case definitions for diabetes in ICD-9-coded and ICD-10-coded data in adult populations. BMJ Open 2016;6:e009952.

37. Liu B, Hadzi-Tosev M, Liu Y, et al. Accuracy of International Classification of Diseases, 10th Revision Codes for Identifying Sepsis: A Systematic Review and Meta-Analysis. Crit Care Explor 2022;4:e0788.

38. Logan R, Davey P, De Souza N, et al. Assessing the accuracy of ICD-10 coding for measuring rates of and mortality from acute kidney injury and the impact of electronic alerts: an observational cohort study. Clin Kidney J 2020;13:1083–1090.

39. O’Malley KJ, Cook KF, Price MD, et al. Measuring diagnoses: ICD code accuracy. Health Serv Res 2005;40:1620–39.

40. Lindner HA, Thiel M, Schneider-Lindner V. Clinical ground truth in machine learning for early sepsis diagnosis. Lancet Digit Health 2023;5:e338–e339.

41. Kaplan DE, Ripoll C, Thiele M, et al. AASLD Practice Guidance on risk stratification and management of portal hypertension and varices in cirrhosis. Hepatology 2024;79:1180–1211.

42. Karvellas CJ, Bajaj JS, Kamath PS, et al. AASLD Practice Guidance on Acute-on-chronic liver failure and the management of critically ill patients with cirrhosis. Hepatology 2024;79:1463–1502.

43. Valenstein PN. Evaluating diagnostic tests with imperfect standards. Am J Clin Pathol 1990;93:252–8.

44. Far AT, Bastani A, Lee A, et al. Evaluating the positive predictive value of code-based identification of cirrhosis and its complications utilizing GPT-4. Hepatology 2024.

