## Supplementary for "Achieving Expert-Level Clinical Infection Detection with LLMs from Clinical Documents: Validation in Complex Patient Cases with Cirrhosis"

### SUPPLEMENTARY INFORMATION

#### Supplementary Method Section 1. Detailed LLM prompt, setup, and output logic.

To extract infection-related information from clinical notes, we utilized Anthropic's Claude 3.5 Sonnet, a 175-billion-parameter LLM with a 200,000-token context window. A zero-shot chain of thought (CoT) prompting approach was employed, with no retrieval-augmented generation (RAG); the complete 72-hour clinical note set was fed directly into the model. To enhance reproducibility and minimize variability in the LLM's responses, the temperature parameter was set to 0.1.

The prompt was designed to function as a clinical reasoning assistant, guiding the LLM through a structured, evidence-based infection assessment process designed to mirror physician decision-making. It specified key decision points and required outputs to ensure interpretability and clinical relevance. The prompt also required the LLM to provide a confidence level for each decision point, serving as a self-reflection mechanism on the sufficiency of textual evidence.

The LLM followed a stepwise, systematic decision process to classify infections. It first evaluated the presence of an acute infection based on objective indicators such as vital signs, laboratory results, imaging findings, microbiological evidence, and provider assessments. Acute infections were defined as those with rapid onset and resolution within days to weeks; chronic infections such as HIV or Hepatitis C were excluded. If an acute infection was identified, the model classified the pathogen type (bacterial, viral, or fungal) and generated concise, evidence-based justifications for each decision. If applicable, infections were further categorized by anatomical site using predefined categories based on established definitions, including lower respiratory tract, urinary tract, intra-abdominal, gastrointestinal, skin and soft tissue, bone and joint, central nervous system, primary bloodstream, catheter-related, neutropenic fever, ear, nose, throat, and reproductive tract infections.

To enhance classification accuracy, the model was explicitly instructed to disregard prophylactic or empiric antibiotic use as the sole criterion for infection. This process incorporated conditional logic, skipping irrelevant downstream questions (e.g., pathogen classification was bypassed if no infection was detected) to enhance efficiency. For each classification (e.g. presence of infection, pathogen type, and infection site), the model assigned a confidence level (strong, intermediate, or low) based on the quality and clarity of supporting documentation. Strong confidence indicated well-documented evidence consistent with typical clinical patterns (e.g., positive cultures, abnormal imaging, explicit provider diagnosis for infection presence; multiple negative cultures, normal inflammatory markers, absence of clinical signs for infection absence). Intermediate confidence reflected partial or inconclusive support (e.g., suspected infection with missing diagnostic elements; mild symptoms despite negative tests). Low confidence was assigned when documentation was ambiguous, conflicting, or insufficient to confirm or rule out infection reliably. This confidence annotation aimed to enhance the interpretability and utility of the model's outputs for downstream applications. The complete prompt is available in **Supplementary Material 2**.

To maintain consistency and facilitate evaluation, the model generated structured JSON output containing binary responses for infection presence and pathogen type, along with infection locations and corresponding justifications. The output also included a stepwise reasoning array that mimics expert clinical reasoning by 1) summarizing key points from documentation, 2) correlating clinical findings with diagnostic data, and 3) explaining the final infection classification. Each output adhered to a predefined schema with specified data types, ensuring consistency and reliability.

**Supplementary Results Section 1**. Prompt development cohort.

**Supplementary Results Section 1.1.** Cohort characteristics.

The *prompt development cohort* consisted of 196 encounters. Of these, 79.6% (n=156) had documented infections or strong clinical evidence of infection within the first 48 hours of admission. The etiology of infection was predominantly bacterial t (n=119, 60.7%), followed by viral (n=31, 15.8%) and fungal (n=17, 8.7%). The most common infection sites were lower respiratory tract (n=49, 25.0%), urinary tract (n=23, 11.7%), skin and soft tissue (n=19, 9.7%), and intra-abdominal region (n=18, 9.2%).

**Supplementary Results Section 1.2.** Infection presence identification results.

INFEHR was evaluated against physician chart review for infection presence. The pipeline achieved an overall accuracy of 94.9%, with a sensitivity of 99.3%, specificity of 83.3%, PPV of 94.0%, and NPV of 97.8%.

The average confidence score assigned by the model was 2.94 (1=Low, 2=Intermediate, 3=Strong), with infection-positive encounters averaging 2.92 and infection-negative encounters averaging 3.0. Intermediate confidence was assigned in 12 encounters (6.1%), of which 5 (41.7%) were false positives (as determined by physician review). Limiting the analysis to cases with strong model confidence (n=184) improved performance: accuracy increased to 97.3%, specificity to 91.8%, and PPV to 97.1%, while sensitivity and NPV remained high at 99.3% and 97.8%, respectively.

An error analysis identified 9 false positives and 1 false negative. Most false positives (n=6, 66.7%) involved empiric antibiotic use or clinical deterioration (e.g., liver failure causing shock) without a clearly documented infection source. In several cases, the model appeared to overinterpret inconclusive findings, such as imaging abnormalities or culture-negative results. One-third of false positives involved infections that were recently treated or resolved, suggesting difficulty distinguishing active from past infections. The single false negative occurred in a patient with recent neutropenia, but no clear evidence of infection documented during the 72-hour review window.

**Supplementary Results Section 1.3.** Pathogen type results.

The performance of INFEHR in classifying infections by pathogen type within the prompt development cohort is detailed in **Supplementary Table 4**. Overall, the model demonstrated high sensitivity across all pathogen types, but PPV varied with infection prevalence.

The average model-assigned confidence scores were 2.74 for bacterial, 2.93 for viral, and 2.95 for fungal infections. When limiting the analysis to cases with strong model confidence, performance improved across all metrics (**Supplementary Table 4**). Accuracy for bacterial, viral, and fungal classifications increased to 96.6%, 97.8%, and 97.3%, respectively, and PPVs rose to 98.8%, 90.9%, and 82.4%.

**Supplementary Results Section 1.4.** Infection location results.

The performance of the INFEHR pipeline in classifying infections by anatomical location within the prompt development cohort is presented in **Supplementary Table 5**. The model demonstrated high accuracy and specificity (>90%) across all infection sites. NPV values were also consistently high, exceeding 97% in most categories. Among common infection sites, sensitivity was highest for lower respiratory tract infections, urinary tract infections, and skin and soft tissue infections. PPV varied more widely, with lower values observed for less common or diagnostically challenging sites such as primary bloodstream infections and gastrointestinal infections.

We further analyzed the relationship between model confidence and PPV across infection site classifications (**Supplementary Table 5**). Because INFEHR only outputs suspected/positive infection sites, reported confidence scores reflect only encounters where a site was identified. Average confidence scores ranged from 2.44 to 3.00. INFEHR assigned the maximum confidence level showed perfect PPV for neutropenic fever and reproductive tract infections. However, this relationship did not consistently hold across all locations. For example, primary bloodstream infections had a relatively high average confidence score (2.81) but the lowest PPV (31.6%), whereas intra-abdominal infections had the lowest average confidence (2.44) yet achieved a higher PPV (68.4%).

**Supplementary Table 1.** List of ICD-10 infection codes used in the evaluation.

| Infection Sites | Pathogen Type | ICD-10 Codes |
|---|---|---|
| Lower Respiratory Tract | Bacterial | A02.22, A15.0, A15.4, A15.5, A15.6, A15.7, A15.8, A15.9, A20.2, A21.2, A22.1, A31.0, A37.00, A37.01, A37.10, A37.11, A37.80, A37.81, A37.90, A37.91, A42.0, A43.0, A48.1, A52.72, J13, J14, J15.0, J15.1, J15.2, J15.211, J15.212, J15.29, J15.3, J15.4, J15.5, J15.61, J15.69, J15.7, J15.8, J15.9, J16.0, J16.8, J18.0, J18.1, J18.8, J18.9, J20.0, J20.1, J20.2, J44.0, J47.0, J85.0, J85.1, J85.2, J85.3, J86.0, J86.9, J98.51, J95.02, O98.011, O98.012, O98.013, O98.019 |
| | Viral | B01.2, B05.2, B25.0, J09.X1, J09.X2, J10.00, J10.01, J10.08, J10.1, J11.00, J11.08, J12.0, J12.1, J12.2, J12.3, J12.81, J12.82, J12.89, J12.9, J20.3, J20.4, J20.5, J20.6, J20.7, J21.0, J21.1, U07.1 |
| | Fungal | B37.1, B38.0, B38.1, B38.2, B39.0, B39.1, B39.2, B44.0, B59 |
| | Non-specific | J20.8, J20.9, J21.8, J21.9, J22, T86.812 |
| Urinary Tract | Bacterial | A18.10, A18.11, A18.12, A18.13, A36.85, A52.75, A54.01, A54.21, A56.01, N01.0, N30.00, N30.01, N30.10, N30.11, N30.20, N30.21, N30.30, N30.31, N30.80, N30.81, N30.90, N30.91, N34.0, N34.2, N34.3, N39.0, N99.511, N99.521, N99.531, O03.38, O03.88, O04.88, O07.38, O08.83, O23.00, O23.01, O23.02, O23.03, O23.10, O23.11, O23.12, O23.13, O23.20, O23.21, O23.22, O23.23, O23.30, O23.31, O23.32, O23.33, O23.40, O23.41, O23.42, O23.43, O86.19, O86.20, O86.21, O86.22, T08.35 |
| | Fungal | B37.41 |
| Intra-abdominal | Bacterial | 0D5J.0Z3, 0D5J.0ZZ, 0D5J.3Z3, 0D5J.3ZZ, 0D5J.4Z3, 0D5J.4ZZ, 0D5J.7ZZ, 0D5J.8ZZ, 0D9J.00Z, 0D9J.0ZX, 0D9J.0ZZ, 0D9J.3ZX, 0D9J.40Z, 0D9J.4ZX, 0D9J.4ZZ, 0D9J.70Z, 0D9J.7ZX, 0D9J.7ZZ, 0D9J.80Z, 0D9J.8ZX, 0D9J.8ZZ, 0DBJ.0ZX, 0DBJ.0ZZ, 0DBJ.3ZX, 0DBJ.3ZZ, 0DBJ.4ZG, 0DBJ.4ZX, 0DBJ.4ZZ, 0DBJ.7ZX, 0DBJ.7ZZ, 0DBJ.8ZX, 0DBJ.8ZZ, 0DCJ.0ZZ, 0DCJ.3ZZ, 0DCJ.4ZZ, 0DCJ.7ZZ, 0DCJ.8ZZ, 0DDJ.3ZX, 0DDJ.4ZX, 0DDJ.8ZX, 0DFJ.0ZZ, 0DFJ.3ZZ, 0DFJ.4ZZ, 0DFJ.7ZZ, 0DFJ.8ZZ, 0DNJ.7ZZ, 0DNJ.8ZZ, 0DQJ.0ZZ, 0DQJ.3ZZ, 0DQJ.4ZZ, 0DQJ.7ZZ, 0DQJ.8ZZ, 0DTJ.0ZZ, 0DTJ.4ZG, 0DTJ.4ZZ, 0DTJ.7ZZ, 0DTJ.8ZZ, A18.31, A18.32, A18.39, A18.83, A21.3, A22.2, A42.1, K25.1, K25.2, K25.5, K25.6, K26.1, K26.2, K26.5, K26.6, K27.1, K27.2, K27.5, K27.6, K28.1, K28.2, K28.5, K28.6, K35.200, K35.201, K35.209, K35.210, K35.211, K35.219, K35.30, K35.31, K35.32, K35.33, K35.890, K35.891, K36, K37, K57.00, K57.12, K57.20, K57.32, K57.40, K57.52, K57.80, K57.92, K61, K63.0, K65.0, K65.1, K65.2, K65.8, K65.9, K67, K68.11, |

| | | |
|---|---|---|
| | | K68.12, K68.19, K75.0, K80.00, K80.01, K80.10, K80.11, K80.12, K80.13, K80.18, K80.19, K80.30, K80.31, K80.32, K80.33, K80.34, K80.35, K80.36, K80.37, K80.40, K80.41, K80.42, K80.43, K80.44, K80.45, K80.46, K80.47, K80.60, K80.61, K80.62, K80.63, K80.64, K80.65, K80.66, K80.67, K81.0, K81.1, K81.2, K81.9, K82.2, K82.A1, K82.A2, K83.09, K85.02, K85.12, K85.22, K85.32, K85.82, K85.92 |
| | Viral | B15.0, B15.9, B16.0, B16.1, B16.2, B16.9, B17.0, B17.10, B17.11, B17.2, B17.8, B25.1, B25.2, B26.81 |
| | Non-specific | T86.43 |
| Gastrointestinal | Bacterial | A02.0, A03.0, A03.1, A03.2, A03.3, A03.8, A03.9, A04.0, A04.1, A04.2, A04.3, A04.4, A04.5, A04.6, A04.71, A04.72, A04.8, A04.9, A05.0, A05.2, A05.3, A05.4, A05.5, A05.8, A05.9 |
| | Viral | A00.0, A00.1, A00.9 |
| | Fungal | B37.81, B37.82 |
| | Non-specific | A08.8, A09, K22.3 |
| Skin and Soft Tissue | Bacterial | A18.4, A22.0, A31.1, A36.3, A42.2, A43.1, A51.31, A51.32, A51.39, B55.1, B55.2, B65.3, B87.1, B87.2, B87.3, B87.4, B87.81, B87.82, B87.89, B87.9, B88.0, B88.1, L00, L01.00, L01.01, L01.02, L01.03, L01.09, L01.1, L02.01, L02.02, L02.03, L02.11, L02.12, L02.13, L02.211, L02.212, L02.213, L02.214, L02.215, L02.216, L02.219, L02.221, L02.222, L02.223, L02.224, L02.225, L02.226, L02.229, L02.231, L02.232, L02.233, L02.234, L02.235, L02.236, L02.239, L02.31, L02.32, L02.33, L02.411, L02.412, L02.413, L02.414, L02.415, L02.416, L02.419, L02.421, L02.422, L02.423, L02.424, L02.425, L02.426, L02.429, L02.431, L02.432, L02.433, L02.434, L02.435, L02.436, L02.439, L02.511, L02.512, L02.519, L02.521, L02.522, L02.529, L02.531, L02.532, L02.539, L02.611, L02.612, L02.619, L02.621, L02.622, L02.629, L02.631, L02.632, L02.639, L02.811, L02.818, L02.821, L02.828, L02.831, L02.838, L02.91, L02.92, L02.93, L03.011, L03.012, L03.019, L03.021, L03.022, L03.029, L03.031, L03.032, L03.039, L03.041, L03.042, L03.049, L03.111, L03.112, L03.113, L03.114, L03.115, L03.116, L03.119, L03.121, L03.122, L03.123, L03.124, L03.125, L03.126, L03.129, L03.211, L03.212, L03.213, L03.221, L03.222, L03.311, L03.312, L03.313, L03.314, L03.315, L03.316, L03.317, L03.319, L03.321, L03.322, L03.323, L03.324, L03.325, L03.326, L03.327, L03.329, L03.811, L03.818, L03.891, L03.898, L03.90, L03.91, L04, L05.01, L05.02, L05.91, L05.92, L08.0, L08.81, L08.82, L08.89, L08.9, M60.000, M60.001, M60.002, M60.003, M60.004, M60.005, M60.009, M60.011, M60.012, M60.019, M60.021, M60.022, M60.029, M60.031, |

| | | |
|---|---|---|
| | | M60.032, M60.039, M60.041, M60.042, M60.043, M60.044, M60.045, M60.046, M60.051, M60.052, M60.059, M60.061, M60.062, M60.069, M60.070, M60.071, M60.072, M60.073, M60.074, M60.075, M60.076, M60.077, M60.078, M60.08, M60.09, M65.0, M65.1, M72.6, N61.0, N61.1, O86.00, O86.01, O86.02, O86.03, O86.09, O91.011, O91.012, O91.013, O91.019, O91.02, O91.03, O91.111, O91.112, O91.113, O91.119, O91.12, O91.13, O91.211, O91.212, O91.213, O91.219, O91.22, O91.23, T81.40XA, T81.41XA, T81.42XA, T81.43XA, T81.49XA |
| | Fungal | B35, B36, B37.2, B38.3 |
| Bone and Joint | Bacterial | A18.02, A54.42, M00.061, M00.062, M00.069, M00.161, M00.162, M00.169, M00.261, M00.262, M00.269, M00.861, M00.862, M00.869, M00.9, M01.X61, M01.X62, M01.X69, M71.061, M71.062, M71.069, M71.161, M71.162, M71.169, M86.061, M86.062, M86.069, M86.161, M86.162, M86.169, M86.261, M86.262, M86.269, M86.361, M86.362, M86.369, M86.461, M86.462, M86.469, M86.561, M86.562, M86.569, M86.661, M86.662, M86.669, M86.8X6, T84.53XA, T84.54XA, T84.620A, T84.621A, T84.622A, T84.623A, T84.624A, T84.625A, T84.629A, T84.7XXA, A02.24, A18.01, A18.03, A51.46, A52.77, A54.41, M46.20, M46.21, M46.22, M46.23, M46.24, M46.25, M46.26, M46.27, M46.28, M46.30, M46.31, M46.32, M46.33, M46.34, M46.35, M46.36, M46.37, M46.38, M46.39, M86.00, M86.011, M86.012, M86.019, M86.021, M86.022, M86.029, M86.031, M86.032, M86.039, M86.041, M86.042, M86.049, M86.051, M86.052, M86.059, M86.061, M86.062, M86.069, M86.071, M86.072, M86.079, M86.08, M86.09, M86.10, M86.111, M86.112, M86.119, M86.121, M86.122, M86.129, M86.131, M86.132, M86.139, M86.141, M86.142, M86.149, M86.151, M86.152, M86.159, M86.161, M86.162, M86.169, M86.171, M86.172, M86.179, M86.18, M86.19, M86.20, M86.211, M86.212, M86.219, M86.221, M86.222, M86.229, M86.231, M86.232, M86.239, M86.241, M86.242, M86.249, M86.251, M86.252, M86.259, M86.261, M86.262, M86.269, M86.271, M86.272, M86.279, M86.28, M86.29, M86.30, M86.311, M86.312, M86.319, M86.321, M86.322, M86.329, M86.331, M86.332, M86.339, M86.341, M86.342, M86.349, M86.351, M86.352, M86.359, M86.361, M86.362, M86.369, M86.371, M86.372, M86.379, M86.38, M86.39, M86.40, M86.411, M86.412, M86.419, M86.421, M86.422, M86.429, M86.431, M86.432, M86.439, M86.441, M86.442, M86.449, M86.451, M86.452, M86.459, M86.461, M86.462, M86.469, M86.471, M86.472, M86.479, M86.48, M86.49, M86.50, M86.511, M86.512, M86.519, M86.521, M86.522, M86.529, M86.531, M86.532, M86.539, M86.541, M86.542, M86.549, M86.551, M86.552, M86.559, M86.561, M86.562, M86.569, M86.571, M86.572, M86.579, M86.58, M86.59, M86.60, M86.611, M86.612, M86.619, M86.621, M86.622, M86.629, M86.631, M86.632, M86.639, M86.641, M86.642, M86.649, M86.651, M86.652, M86.659, M86.661, M86.662, M86.669, M86.671, M86.672, M86.679, M86.68, M86.69, M86.8X0, M86.8X1, M86.8X2, M86.8X3, M86.8X4, M86.8X5, M86.8X6, M86.8X7, |

| | | |
|---|---|---|
| | | M86.8X8, M86.8X9, M86.9, A02.23, A18.02, A18.09, A39.83, A39.84, A54.40, A54.42, A54.43, A54.49, A66.6, M00.00, M00.011, M00.012, M00.019, M00.021, M00.022, M00.029, M00.031, M00.032, M00.039, M00.041, M00.042, M00.049, M00.051, M00.052, M00.059, M00.061, M00.062, M00.069, M00.071, M00.072, M00.079, M00.08, M00.09, M00.10, M00.111, M00.112, M00.119, M00.121, M00.122, M00.129, M00.131, M00.132, M00.139, M00.141, M00.142, M00.149, M00.151, M00.152, M00.159, M00.161, M00.162, M00.169, M00.171, M00.172, M00.179, M00.18, M00.19, M00.20, M00.211, M00.212, M00.219, M00.221, M00.222, M00.229, M00.231, M00.232, M00.239, M00.241, M00.242, M00.249, M00.251, M00.252, M00.259, M00.261, M00.262, M00.269, M00.271, M00.272, M00.279, M00.28, M00.29, M00.80, M00.811, M00.812, M00.819, M00.821, M00.822, M00.829, M00.831, M00.832, M00.839, M00.841, M00.842, M00.849, M00.851, M00.852, M00.859, M00.861, M00.862, M00.869, M00.871, M00.872, M00.879, M00.88, M00.89, M00.9, M01.X0, M01.X11, M01.X12, M01.X19, M01.X21, M01.X22, M01.X29, M01.X31, M01.X32, M01.X39, M01.X41, M01.X42, M01.X49, M01.X51, M01.X52, M01.X59, M01.X61, M01.X62, M01.X69, M01.X71, M01.X72, M01.X79, M01.X8, M01.X9 |
| Central Nervous System | Bacterial | A17.0, A27.81, A27.89, A39.0, A50.40, A50.41, A50.42, A50.43, A50.45, A50.49, A51.41, A52.13, A52.14, A52.2, A54.81, G00, G01, G06, G07 |
| | Viral | A80.0, A80.1, A80.2, A80.30, A80.39, A80.9, A81, A82.0, A82.1, A82.9, A83.0, A83.1, A83.2, A83.3, A83.4, A83.5, A83.6, A83.8, A83.9, A84.0, A84.1, A84.81, A84.89, A84.9, A85.0, A85.1, A85.2, A85.8, A86, A87.0, A87.1, A87.2, A87.8, A87.9, A88.0, A88.8, A89, A92.2, B00.3, B00.4, B00.82, B01.11, B01.12, B02.1, B02.24, B04.0, B05.0, B06.01, B06.02, B06.09, B10.01, B10.09, B26.1, B26.2, G03.2 |
| | Fungal | B37.5, B38.4, B45.1 |
| Primary Bloodstream | Bacterial | A39.51, A52.03, I33.0, I33.9, T80.22XA, T80.29XA |
| | Viral | B33.21 |
| | Fungal | B37.6, B37.7 |
| | Non-specific | I38, I39 |
| Catheter-related | Non-specific | T80.211A, T80.212A, T80.218A, T80.219A |
| Neutropenic Fever | Non-specific | D70.3 |

| | | |
|---|---|---|
| Ear, Nose, Throat | Bacterial | A18.6, A36.0, A36.1, A36.2, A54.5, A56.4, A69.0, H60.00, H60.01, H60.02, H60.03, H60.10, H60.11, H60.12, H60.13, H60.20, H60.21, H60.22, H60.23, H60.311, H60.312, H60.313, H60.319, H60.321, H60.322, H60.323, H60.329, H60.331, H60.332, H60.333, H60.339, H60.391, H60.392, H60.393, H60.399, H60.511, H60.512, H60.513, H60.519, H60.90, H60.91, H60.92, H60.93, H61.001, H61.002, H61.003, H61.009, H65.00, H65.01, H65.02, H65.03, H65.04, H65.05, H65.06, H65.07, H65.191, H65.192, H65.193, H65.194, H65.195, H65.196, H65.197, H65.199, H66.001, H66.002, H66.003, H66.004, H66.005, H66.006, H66.007, H66.009, H66.011, H66.012, H66.013, H66.014, H66.015, H66.016, H66.017, H66.019, H66.10, H66.11, H66.12, H66.13, H66.20, H66.21, H66.22, H66.23, H66.3X1, H66.3X2, H66.3X3, H66.3X9, H66.40, H66.41, H66.42, H66.43, H66.90, H66.91, H66.92, H66.93, H67.1, H67.2, H67.3, H67.9, H68.001, H68.002, H68.003, H68.009, H68.011, H68.012, H68.013, H68.019, H70.001, H70.002, H70.003, H70.009, H70.011, H70.012, H70.013, H70.019, H70.091, H70.092, H70.093, H70.099, H70.10, H70.11, H70.12, H70.13, H70.891, H70.892, H70.893, H70.899, H75.00, H75.01, H75.02, H75.03, J01.00, J01.01, J01.10, J01.11, J01.20, J01.21, J01.30, J01.31, J01.40, J01.41, J01.80, J01.81, J01.90, J01.91, J02.0, J02.8, J02.9, J03.00, J03.01, J03.80, J03.81, J03.90, J03.91, J04.0, J04.2, J04.30, J04.31, J05.0, J05.10, J05.11, J36, J37.1, J39.0, K12.2 |
| | Viral | B00.1, B00.2, B05.3, B08.5, J00, J11.1 |
| | Fungal | B37.0, B37.83, B37.84 |
| | Non-specific | H73.001, H73.002, H73.003, H73.009, H73.011, H73.012, H73.013, H73.019, H73.091, H73.092, H73.093, H73.099 |
| Reproductive Tract | Bacterial | A18.14, A18.15, A18.16, A18.17, A18.18, A51.0, A54.00, A54.01, A54.02, A54.03, A54.09, A54.21, A54.22, A54.23, A54.24, A54.29, A55, A56.00, A56.01, A56.02, A56.09, A56.11, A56.19, A56.2, A56.8, A57, A58, N39.80, N41.0, N41.2, N41.3, N43.1, N48.1, N48.21, N48.22, N49.3, N70.01, N70.02, N70.03, N70.11, N70.12, N70.13, N70.91, N70.92, N70.93, N73.0, N73.1, N73.2, N73.3, N73.4, N73.5, N73.8, N73.9, N74, N75.1, N76.4, N76.82, N94.810, N98.0, O03.0, O03.38, O03.88, O04.5, O04.88, O07.0, O07.38, O08.0, O08.83, O23.00, O23.01, O23.02, O23.03, O23.10, O23.11, O23.12, O23.13, O23.20, O23.21, O23.22, O23.23, O23.30, O23.31, O23.32, O23.33, O23.40, O23.41, O23.42, O23.43, O23.511, O23.512, O23.513, O23.519, O23.521, O23.522, O23.523, O23.529, O23.591, O23.592, O23.593, O23.599, O23.90, O23.91, O23.92, O23.93, O41.1010, O41.1011, O41.1012, O41.1013, O41.1014, O41.1015, O41.1019, O41.1020, O41.1021, O41.1022, O41.1023, O41.1024, O41.1025, O41.1029, O41.1030, O41.1031, O41.1032, O41.1033, O41.1034, O41.1035, O41.1039, O41.1090, O41.1091, O41.1092, O41.1093, O41.1094, O41.1095, O41.1099, O41.1210, O41.1211, O41.1212, |

| | | |
|---|---|---|
| | | O41.1213, O41.1214, O41.1215, O41.1219, O41.1220, O41.1221, O41.1222, O41.1223, O41.1224, O41.1225, O41.1229, O41.1230, O41.1231, O41.1232, O41.1233, O41.1234, O41.1235, O41.1239, O41.1290, O41.1291, O41.1292, O41.1293, O41.1294, O41.1295, O41.1299, O41.1410, O41.1411, O41.1412, O41.1413, O41.1414, O41.1415, O41.1419, O41.1420, O41.1421, O41.1422, O41.1423, O41.1424, O41.1425, O41.1429, O41.1430, O41.1431, O41.1432, O41.1433, O41.1434, O41.1435, O41.1439, O41.1490, O41.1491, O41.1492, O41.1493, O41.1494, O41.1495, O41.1499, O86.11, O86.12, O86.13, O86.29, O86.89, O98.111, O98.112, O98.113, O98.119, O98.211, O98.212, O98.213, O98.219 |
| | Viral | A60.00, A60.01, A60.02, A60.03, A60.04, A60.09, A60.1, A60.9, B26.0 |
| | Fungal | B37.31, B37.32, B37.42, B37.49 |
| | Non-specific | A63.8, A64, O98.33, O98.83, O98.93, O98.311, O98.312, O98.313, O98.319 |

**Supplementary Table 2.** Characteristics of the prompt development and the evaluation cohorts.

| Characteristics | Prompt Development | Evaluation |
|---|---|---|
| Number of encounters | 196 | 1000 |
| Unique individuals | 180 | 736 |
| Age at visit (yrs) (median, 25%- 75% percentile) | 59.0 (51.7 - 65.3) | 59.0 (51.9 - 66.1) |
| % Female | 34.7% | 37.5% |
| Length of stay in days (median, 25%- 75% percentile) | 6.6 (3.7, 13.4) | 6.8 (4.0, 12.0) |
| % ICU admission | 15.3% | 20.8% |
| % In-hospital Mortality | 3.6% | 6.9% |
| % Infection | 79.6% | 59.7% |
| % Multiple Pathogen Type | 12.8% | 4.8% |
| % Multiple Infective Sites | 14.5% | 11.2% |

**Supplementary Table 3.** Adjudicated correctness of INFEHR infection presence classification in discordant cases (false positives and false negatives), stratified by initial physician and INFEHR confidence levels. Values indicate the number of adjudicated cases where INFEHR was ultimately deemed correct. Percentages indicate the proportion of cases within that specific confidence stratum (row/column intersection) where INFEHR was correct.

| **Confidence Level** | | **INFEHR** | |
|---|---|---|---|
| | | **Strong** | **Intermediate/Low** |
| **Physician** | **Strong** | 42 (67.7%) | 6 (25.0%) |
| | **Low** | 25 (78.1%) | 9 (52.9%) |

**Supplementary Table 4.** INFEHR performance for pathogen type classification in the prompt development cohort: overall vs. strong-confidence subgroup.

| **Cohort** | **Pathogen Type** | **N** | **Accuracy** | **Sensitivity** | **Specificity** | **PPV** | **NPV** |
|---|---|---|---|---|---|---|---|
| All | Bacterial | 196 | 0.908 | 0.916 | 0.896 | 0.932 | 0.873 |
| | Viral | 196 | 0.959 | 0.968 | 0.958 | 0.811 | 0.994 |
| | Fungal | 196 | 0.949 | 0.882 | 0.955 | 0.652 | 0.988 |
| Strong Confidence Only | Bacterial | 148 | 0.966 | 0.953 | 0.984 | 0.988 | 0.939 |
| | Viral | 183 | 0.978 | 0.968 | 0.980 | 0.909 | 0.993 |
| | Fungal | 186 | 0.973 | 0.875 | 0.982 | 0.824 | 0.988 |

**Supplementary Table 5.** INFEHR performance for infection location classification in the prompt development cohort.

| Infection Location | TP | TN | FP | FN | Accuracy | Sensitivity | Specificity | PPV | NPV | Confidence |
|---|---|---|---|---|---|---|---|---|---|---|
| Lower Respiratory Tract | 39 | 138 | 9 | 10 | 0.913 | 0.918 | 0.912 | 0.776 | 0.971 | 2.50 |
| Urinary Tract | 19 | 169 | 4 | 4 | 0.964 | 0.913 | 0.971 | 0.808 | 0.988 | 2.70 |
| Intra-abdominal | 13 | 175 | 3 | 5 | 0.944 | 0.722 | 0.966 | 0.684 | 0.972 | 2.44 |
| Gastrointestinal | 7 | 185 | 2 | 2 | 0.964 | 0.667 | 0.979 | 0.600 | 0.984 | 2.67 |
| Skin and Soft Tissue | 17 | 172 | 5 | 2 | 0.969 | 0.895 | 0.977 | 0.810 | 0.989 | 2.91 |
| Bone and Joint | 6 | 184 | 5 | 1 | 0.974 | 0.857 | 0.979 | 0.600 | 0.995 | 2.64 |
| Central Nervous System | 6 | 188 | 1 | 1 | 0.995 | 0.857 | 1.000 | 1.000 | 0.995 | 2.86 |
| Primary Bloodstream | 7 | 172 | 14 | 3 | 0.913 | 0.600 | 0.930 | 0.316 | 0.977 | 2.81 |
| Catheter-related | 3 | 188 | 5 | 0 | 0.985 | 1.000 | 0.985 | 0.500 | 1.000 | 2.87 |
| Neutropenic Fever | 9 | 186 | 0 | 1 | 0.995 | 0.900 | 1.000 | 1.000 | 0.995 | 3.00 |
| Ear, Nose, Throat | 7 | 183 | 4 | 2 | 0.969 | 0.667 | 0.984 | 0.667 | 0.984 | 2.82 |
| Reproductive Tract | 3 | 191 | 0 | 2 | 0.985 | 0.400 | 1.000 | 1.000 | 0.985 | 3.00 |

**Supplementary Table 6.** Performance metrics for ICD-10 code-based infection location classification in the evaluation cohort.

| Infection Location | TP | TN | FP | FN | Accuracy | Sensitivity | Specificity | PPV | NPV |
|---|---|---|---|---|---|---|---|---|---|
| Lower Respiratory Tract | 42 | 837 | 9 | 112 | 0.879 | 0.273 | 0.989 | 0.824 | 0.882 |
| Urinary Tract | 19 | 886 | 6 | 89 | 0.905 | 0.176 | 0.993 | 0.760 | 0.909 |
| Intra-abdominal | 33 | 840 | 15 | 112 | 0.873 | 0.228 | 0.982 | 0.688 | 0.882 |
| Gastrointestinal | 14 | 926 | 3 | 57 | 0.940 | 0.197 | 0.997 | 0.824 | 0.942 |
| Skin and Soft Tissue | 45 | 867 | 11 | 77 | 0.912 | 0.369 | 0.987 | 0.804 | 0.918 |
| Bone and Joint | 11 | 975 | 4 | 10 | 0.986 | 0.524 | 0.996 | 0.733 | 0.990 |
| Central Nervous System | 1 | 991 | 1 | 7 | 0.992 | 0.125 | 0.999 | 0.500 | 0.993 |
| Primary Bloodstream | 2 | 887 | 3 | 108 | 0.889 | 0.018 | 0.997 | 0.400 | 0.891 |
| Catheter-related | 1 | 995 | 0 | 4 | 0.996 | 0.200 | 1.000 | 1.000 | 0.996 |
| Neutropenic Fever | 0 | 996 | 0 | 4 | 0.996 | 0.000 | 1.000 | 0.000 | 0.996 |
| Ear, Nose, Throat | 0 | 986 | 4 | 10 | 0.986 | 0.000 | 0.996 | 0.000 | 0.990 |
| Reproductive Tract | 0 | 995 | 4 | 1 | 0.995 | 0.000 | 0.996 | 0.000 | 0.999 |

**Supplementary Material 1.** Filled TRIPOD-LLM checklist.

**Supplementary Material 2.** Detailed prompt used in the INFEHR pipeline.

You are a clinical reasoning assistant. You will be provided with excerpts from a hospitalized patient's medical documentation from the first 72 hours of admission (including Emergency Department [ED] notes, History & Physical [H&P], and any early progress notes). Your task is to determine if the patient has actual, documented evidence of an acute or active infection—specifically bacterial, fungal, or viral.

Key Points to Consider

1. Acute or active infection often presents with objective findings such as:

- Fever or hypothermia
- Elevated white blood cell count or other inflammatory markers
- Imaging or exam findings suggesting infection (e.g., consolidation on CXR for pneumonia)
- Positive cultures or rapid antigen tests
- Clinical suspicion documented by providers

2. Prophylactic or empiric antibiotics do not automatically confirm an infection. Look for actual supporting evidence:

- Negative cultures plus clinical improvement without targeted antimicrobial therapy indicate no active infection unless there is overwhelming alternative diagnostic evidence.
- Imaging abnormalities alone (e.g., infiltrates on CXR) should not be the sole determinant of infection if cultures or antigen tests are negative and no strong clinical evidence supports infection.
- Mild lab abnormalities or low-grade fevers can have alternative explanations (e.g., autoimmune conditions, post-surgical response, medication effects, trauma-related inflammation, cancer, or chronic disease processes).

- Do NOT assume infection based on suspicion, preliminary assessments, inflammation, or unclear clinical impressions.

3. If the patient is actively receiving treatment for an infection at the time of hospital admission (e.g., currently on antibiotics, antifungals, or antivirals for a diagnosed infection), treat the infection presence as “Yes”. The ongoing treatment indicates an active infectious process.

4. Distinguish between:

- Bacterial infections requiring antibiotic therapy
- Fungal infections (e.g., Candida species, Cryptococcus species, Aspergillus species, Endemic Mycoses, Mucormycosis(Rhizopus species, Mucor species, Rhizomucor species, Lichtheimia species, Cunninghamella species, Apophysomyces species), Pneumocystis)
- Viral infections (e.g., COVID-19, influenza, Parainfluenza, RSV, Adenovirus, or other respiratory viruses, Herpesviruses, Gastrointestinal viruses, Hepatitis A and E, Parvovirus, Measles, Mumps, Rubella)
- By default, classify viral and fungal infections as “No” unless there is clear, supporting laboratory data or explicit clinician documentation confirming the infection.

5. For each decision, assign a confidence level based on the available documentation to assess certainty in both diagnosing and ruling out infection presence, type, and classification.

- Set confidence to Strong: The evidence is clear, well-documented, and aligns with typical clinical presentations. If infection is present, diagnostic tests, imaging, or provider assessments provide a strong basis for classification. If infection is absent, documentation strongly supports ruling out infection (e.g., multiple negative cultures, normal inflammatory markers, no clinical signs).

- Set confidence to Intermediate: There is some supported documentation, but it lacks strong evidence. Infection is suspected, but some key tests or assessments are missing. If infection is absent, documentation suggests it but lacks definitive evidence (e.g., negative cultures but mild symptoms persist).
- Set confidence to Low: The documentation is ambiguous, conflicting, or incomplete. Infection cannot be confirmed or ruled out with certainty due to limited documentation.

Instructions

1. Review all details provided in the <content> tags carefully. Summarize relevant evidence (e.g., fever, imaging, cultures, lab results, provider assessments, signs/symptoms).
2. Answer the following questions in sequential order with the infection_identification tool, using concise, evidence-based reasoning. For each response, include a confidence assessment indicating whether the decision is well-supported by the provided documentation.

(a) Was there an acute or active infection present?

- Yes/No. If "Yes," briefly state the reason or evidence. If "No," explain why not (e.g., negative cultures, no clinical signs, prophylactic antibiotics only).
- If inflammation is present but cultures are negative, do not assume infection—consider alternative explanations.
- Confidence: Strong / Intermediate / Low

(b) If Yes to (a), was there an acute bacterial infection present?

- Yes/No. Provide a short justification (based on clinical, cultures, lab, imaging, etc.).
- Confidence: Strong / Intermediate / Low

(c) If Yes to (a), was there an acute fungal infection present?

- Yes/No. Default to "No" unless there is clear evidence. If "Yes," name the suspected fungus (if known) and the supporting data (positive fungal culture or explicit clinician documentation confirming a fungal infection).
- Confidence: Strong / Intermediate / Low

(d) If Yes to (a), was there an acute viral infection present?

- Yes/No. Default to "No" unless there is clear evidence. If "Yes," specify the virus if known (COVID-19, influenza, RSV, etc.) and any supporting evidence (positive PCR test, antigen test, or explicit clinician documentation confirming a viral infection).
- Confidence: Strong / Intermediate / Low

(e) Type of Infection (if applicable)

- If you answered "Yes" to (a) (and specifically to (b) for bacterial), please classify the most likely infection type from the list below:

1. Lower respiratory tract infection (e.g., pneumonia, parapneumonic effusion, empyema)
2. Urinary tract infection (e.g., cystitis, pyelonephritis, urosepsis)
3. Intra-abdominal infection (e.g., appendicitis, diverticulitis, intra-abdominal abscess, cholangitis)
4. Gastrointestinal infection (e.g., enteric pathogens, C. difficile)
5. Skin and soft tissue infection (e.g., cellulitis, erysipelas, necrotizing fasciitis, abscess)
6. Bone and joint infection (e.g., septic arthritis, osteomyelitis)
7. Central nervous system infection (e.g., meningitis, brain abscess, epidural abscess)
8. Primary bloodstream infection (bacteremia of unclear source, endocarditis, infected vascular graft)
9. Catheter-related infection (e.g., PICC line or dialysis catheter infection)

10. Neutropenic fever (fever in a patient with documented neutropenia, likely bacterial)
11. Ear, nose, throat infection (e.g., sinusitis, otitis media, pharyngitis)
12. Reproductive tract infection (e.g., STI, tubo-ovarian abscess, epididymitis, salpingitis)

3. Provide your reasoning in a clear, stepwise manner to support your conclusions.

Suggested JSON Output Format

Below is an example of how you might structure your response in JSON with the infection_identification tool:


```
{
  "infection_identification": {
    "a_acute_infection_present": {
      "answer": "Yes or No",
      "confidence": "Strong / Intermediate / Low",
      "explanation": "Brief rationale"
    },
    "b_acute_bacterial_infection_present": {
      "answer": "Yes or No",
      "confidence": "Strong / Intermediate / Low",
      "explanation": "Brief rationale"
    },
    "c_acute_fungal_infection_present": {
      "answer": "Yes or No",
      "confidence": "Strong / Intermediate / Low",
```

```
      "explanation": "Brief rationale"
    },
    "d_acute_viral_infection_present": {
      "answer": "Yes or No",
      "confidence": "Strong / Intermediate / Low",
      "explanation": "Brief rationale"
    },
    "e_infection_type": {
      "classification": "One category from the list or 'N/A'",
      "confidence": "Strong / Intermediate / Low",
      "explanation": "Brief rationale (imaging, labs, provider note, etc.)"
    },
    "reasoning_steps": [
      "Step 1: Summarize key points from documentation.",
      "Step 2: Correlate clinical findings with diagnostic data.",
      "Step 3: Explain final conclusion about infection status."
    ]
  }
}
```

How to Use This Format

- Fill each answer field with Yes or No (plus minimal text if needed).
- If you answer “No” to (a), then fields (b), (c), (d), and (e) can remain “No” or have no relevant details, since there is no acute infection.

- If you answer “Yes” to any item, provide brief supporting evidence in the explanation.
- Each decision (a-e) must include a confidence level (“Strong,” “Intermediate,” or “Low”).
- If a confidence level is “Intermediate” or “Low”, the explanation should also clarify why the decision is uncertain.
- The reasoning_steps array should reflect how you arrived at your conclusion step by step (brief bullet points are sufficient).
